# Beta-Thalassemia minor and SARS-CoV-2, prevalence, severity, morbidity and mortality: a systematic review study

**DOI:** 10.1101/2022.07.25.22278009

**Authors:** Edouard Lansiaux, Emmanuel Drouin, Carsten Bolm

**Author notes:** Corresponding authors Additional Author Informations Edouard Lansiaux, Emmanuel Drouin, Carsten Bolm. <u>Funding</u>: There was no specific funding for this manuscript. <u>Author’s contributions statement</u>: EL designed the study and wrote the first draft. All authors revised and accepted the final version of the manuscript.

## Abstract

**Background:** Since the first year of the COVID-19 global pandemic, a hypothesis concerning the possible protection/immunity of beta-thalassemia carriers remains in abeyance.

**Methods:** Three databases (Pubmed Central, Scopus and Google Scholar) were screened and checked in order to extract all studies about incidence of confirmed COVID-19 cases OR mortality rate OR severity assessment OR ICU admission among patients with beta-thalassemia minor, were included in this analysis. The language was limited to English. Studies such as case reports, review studies, and studies that did not have complete data for calculating incidences were excluded.

**Results and discussion:** Three studies upon 2265 were selected. According to our systematic-review meta-analysis, beta-thalassemia carriers could be less COVID-19 affected than general population [IRR= 0.9250(0.5752;1.4877)], affected by COVID-19 with a worst severity [OR=1.5933(0.4884;5.1981)], less admissible into ICU [IRR=0.3620(0.0025;51.6821)] and more susceptible to die from COVID-19 or one of its consequences [IRR=1.8542(0.7819;4.3970)]. However, all of those results stay insignificant with a bad p-value (respectively 0.7479, 0.4400, 0.6881, 0.1610). Other large case-control or registry studies are needed to confirm these trends.

## Introduction

The world has had to face a COVID-19 pandemic for two years and then SARS-CoV-2 variants successive waves. This infection was first discovered in December 2019 in the South China Seafood Market Hubei Province, China [1]. Since the pandemic first months, an epidemiological particularity has been observed concerning minor beta-thalassemia patients: they seem to be protected against severe COVID-19 forms and in this way, the beta-thalassemic trait would confer a certain immunization against COVID-19 [2,3]. Few times after, a scientific niche debate (ed. between experts) was opened on the question: some epidemiological datasets supported this hypothesis [4-7] despite the fact that in vitro experiments tend to demonstrate a lack of a mechanistic link between Hb variants and disease effects [8]. This hypothesis is currently developed on the physiopathological side and based upon 3 main ways: iron, heme and redox metabolism; erythropoietin and ACE cascade; and BCL11A gene.

### Beta-thalassemia minor

Beta-thalassemia is an heterozygous disease caused by a genetic point mutation on a single allele of the □ globin heme on the chromosome 11. Therefore, contrary to the normal adult hemoglobin (96% HbA with □□□□ composition, 2% HbA_2_ with □□□□ composition, 2% HbF with □□□□ composition), patients with beta-thalassemia minor present a special HbA composition with □□□□^e^ or □□□□^+^ model. In this way, they suffer from a mild decrease of □-chain production relative to □-chain and so, a decrease of □-chain available for HbA synthesis. In order to compensate for this default, the human system increases the production of non-affected globins such as HbA2 and HbF [4]. Due to that, a lot of biological cascades are put into game: iron metabolism (due to the iron excess caused by abnormally shaped erythrocytes destruction), ineffective erythropoiesis, free □-chains precipitations…

### Iron, Heme and Redox

The transcribed, non-structural SARS-CoV-2 proteins ORF 8, 3a, and 10 lead critical roles during the SARS-CoV-2 replication and COVID-19 pathogenesis (by activating NF-kB mediated inflammation and immune responses)[9]. The initial hypothesis (i.e. beta-thalassemia minor protection against COVID-19) was based upon a preprint which showed that SARS-CoV-2 ORF8 and surface glycoproteins could gather with the porphyrin in order to form a complex. Meanwhile, ORF1ab, ORF10, and ORF3a proteins could dissociate the iron ions from the heme (viral proteins may interact with α-chains via the Isd domains and then break β-chains using the IgA1 protease structure) by attacking the heme (porphyrin which is initially formed into the mitochondria) on the hemoglobin 1-beta chain [9].

#### Beta-thalassemia patients involve three cell protection systems during erythropoiesis

In β-thalassemia, iron control (and therefore the control of heme) is a priority for cell survival in order to avoid the overload or the proteotoxicity caused by the iron deficiency. In this last scenario (lack of iron), heme-regulated inhibitor of protein translation (HRI) allows to inhibit globin translation into heme-deficient erythroid precursors [10]; and that thanks to the heme-regulated eukaryotic initiation factor-2α (eIF2α) kinase which phosphorylates eIF2 subunit. According to *in vitro* studies, the activation of HRI (with ROS implication) needs the heat shock proteins 70 and 90 presence [11]. An *in vivo* experiment (beta-thalassemic animal models with a genetic deficiency of HRI have shown a more severe hematological phenotype than normal β-thalassemia ones) supports the eIF2α main role in stress erythropoiesis [12]. The activating transcription factor 4 (ATF4) mRNA translational up-regulation by the HRI-eIF2αP signaling pathway was required to soften proteotoxicity and support the differentiation of erythroids [13] (Figure 1.A.). Therefore, the HRI limited proteotoxicity and permitted the ATF4 protein expression in order to keep main mitochondrial functions and oxidative homeostasis [14] (Figure 1.A.).

**Figure 1.A.**
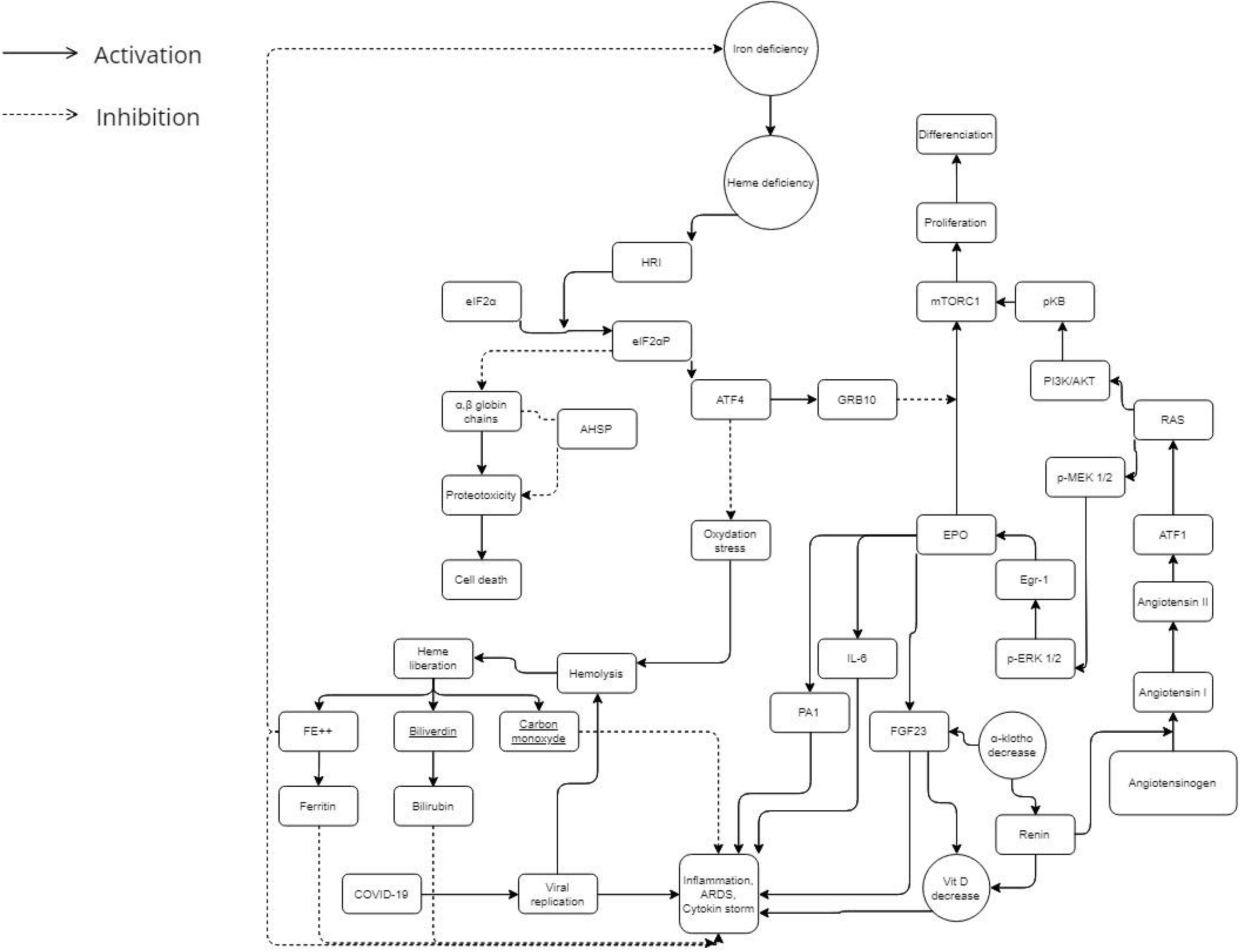
Iron, angiotensin and erythropoietin cascades during COVID-19 infection in a normal metabolism. Into the general population, the metabolic balance leads to a more or less severe possible COVID-19 infection.

The heme consumption has a certain importance in erythropoiesis (and especially when you have an overload of iron due to beta-thalassemia). HO-1 (heme oxygenase-1) is a protein which catalyzes heme catabolism [15]. It is generally considered as a protective enzyme due to its pro-oxidant “free” heme breakdown ability and release of antioxidants such as biliverdin and bilirubin (Figures 1.A. and 1.B.). Several studies demonstrated that some HO-1 gene polymorphisms, especially the promoter region GT dinucleotide repeat mutation, controls the inducibility of HO-1 to ROS [16-21]. According to them, GT sequences with short alleles are associated with HO-1 inducibility resulting in an amplified cytoprotection [21]. COVID-19 complications affected patients may have longer GT sequences and decreased vessel hemostasis.

**Figure 1.B.**
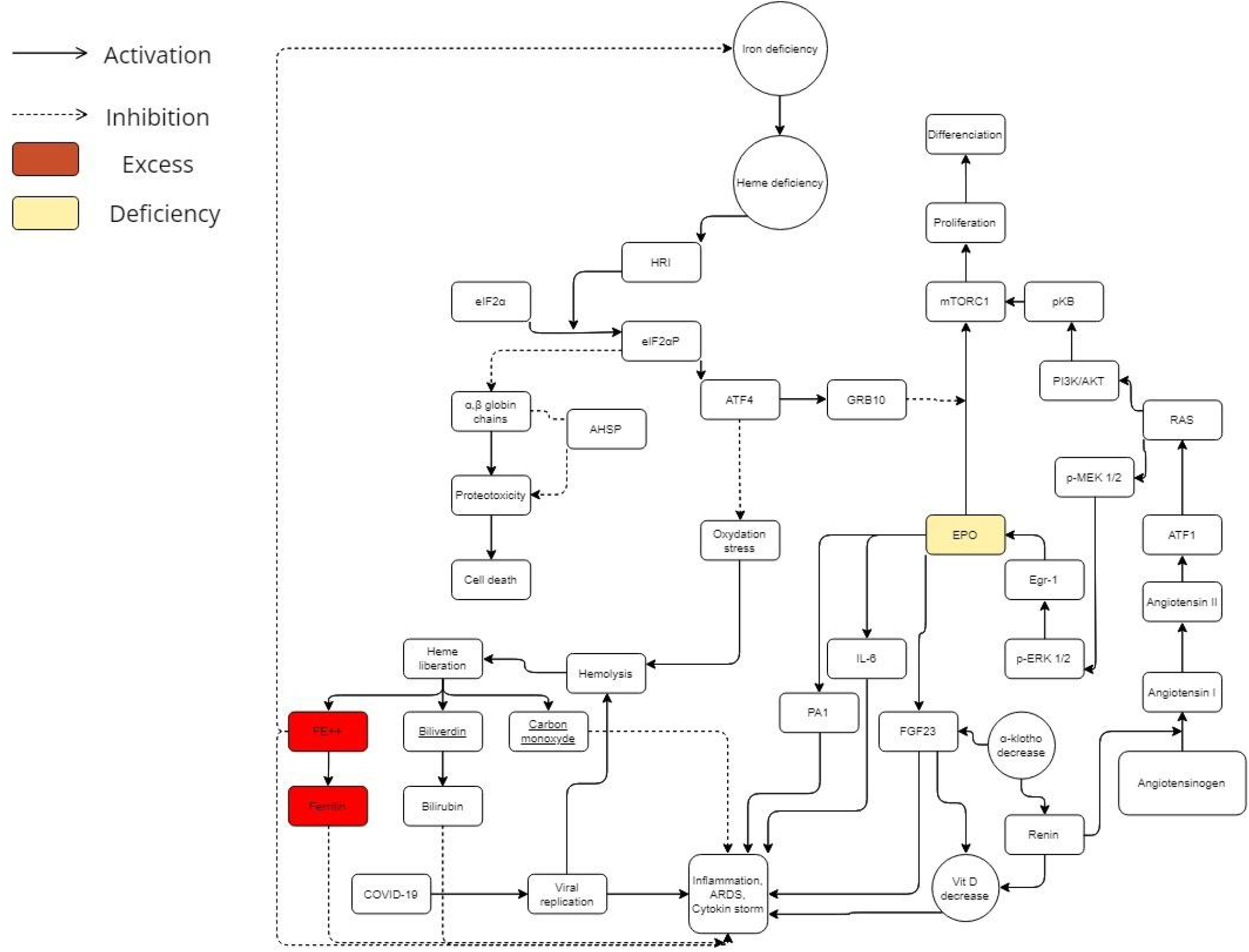
Iron and angiotensin and erythropoietin cascades during COVID-19 infection in a beta-thalassemic carrier metabolism. Beta-thalassemia minor patients have low EPO levels. Furthermore, they have an excess of iron and ferritin. In this way, beta-thalassemia carriers have a different metabolic balance which could explain mild or less severe COVID-19 infection in this population.

Among atypical chaperone systems, the α hemoglobin stabilizing protein (AHSP) plays an important role in the beta-thalassemic erythropoiesis. AHSP coheres to α-Hb, averts its precipitation (which induces splenomegaly into the beta-thalassemia minor [9]), and avoids free α-Hb toxicities. The α-Hb attached to AHSP is stronger to the precipitation induced by oxydation [22]. In addition to that, AHSP knock-out animal models displayed semiologic features and an ineffective degree of erythropoiesis related to β-thalassemia [23].

In view of the facts that beta-thalassemia subjects (and more particularly beta-thalassemia minor) do not seem to have a porphyrin deficiency [24,25], and ferritin and iron excess appears to be a risk factor for COVID-19 exacerbation [36], the possible protection/immunization of beta-thalassemic minor patients could be attributed to another physiopathological mechanism than the heme one: absence of the ORFs interaction with the beta-chain hemoglobin (due to its absence in beta-thalassemia). Those beta-thalassemia protective mechanisms previously described might allow to keep a normal rate of porphyrins, iron and ferritin during COVID-19 infection (Figure 1.B.). It may even be that the latter contribute to reducing the cytokine storm by maintaining a redox homeostasis.

### Erythropoietin and ACE way

The symptoms of altitude sickness at high altitude are similar to COVID-19 ones [29]. Erythropoietin (EPO) seems to be the common denominator between COVID-19, beta-thalassemia and altitude sickness [30]. It has been hypothesized that modulations of the host immune system induced by malarial selection pressure thanks to thalassemia/HbE mutations might give this protection as an antimalarial effect [31]. The I/D polymorphism of D allele of angiotensin converting enzyme (ACE) is significantly linked to mild malaria vs severe malaria, that rises ACE levels and subsequently increases angiotensin II (Ang II) production compared to the I allele [32-33]. Therefore, a common denominator needs to be traced in order to demonstrate those two genetic determinants emergence forced by malarial evolutionary pressure. Both genetic determinants (evolutionary thalassemias selection and the ACE D allele) appear to elicit and sustain a potential phylogenetically preserved ancestral protective innate immune response mechanism against pathogen invasion mediated either due to systemic or/and local increases in erythropoietin (EPO) production [34]. In this way, beta-thalassemic patients have low EPO levels according to the degree of anemia [35,36] (Figure 1.B.). Furthermore, concerning the ACE, the increase of activating transcription factor 1 (ATF1) may be an important key of severe COVID-19 explanation [37].

### BCL11A gene possible imputation

The broad variation in COVID-19 infection rates and its severity could potentially be explained by genetic variability between human and animal hosts. In this way, the observed protection/immunization of beta-thalassemia minor against COVID-19 could have a genetic origin: B-cell lymphoma 11 A (BCL11A) gene which was identified thanks to genome-wide association studies (GWAS) as an associated gene with fetal hemoglobin (HbF) production [38] and therefore as a modulator of beta-thalassemia severity [39-41].

However, there are also epidemiological studies which support this hypothesis [42,43] and there is no evidence of a significant relationship between COVID-19 infection and the oxy-hemoglobin dissociation curve [44]. The main purpose of our manuscript is to do an epidemiological systematic-review meta-analysis on this subject in order to support (or not) the possible beta-thalassemia minor protection against COVID-19 (and its severe forms). In this systematic review study, we wanted to determine the prevalence, severity, the ICU admission and mortality rate of COVID-19 in beta-thalassemic minor patients.

## Methods

This systematic review and meta-analysis has been done according to the preferred reporting items for systematic reviews and meta-analyses (PRISMA) checklist [45]. The review was registered at International Prospective Register of Systematic Reviews (PROSPERO) with the registration number CRD42022345452.

### Searched databases

Searches were conducted in PubMed (www.ncbi.nlm.nih.gov/), Elsevier (www.elsevier.com) and Google Scholar (www.scholar.google.com) from 11 July to 17 July 2022, to identify original research articles, reviews and case reports that described β-thalassemic minor patients affected by coronavirus. The screening period for the studies would be from January 2020 to May 2022. Combinations of the following medical subject headings (MeSH), terms and keywords, were used to conduct a comprehensive literature search: (beta-thalassemia OR Cooley’s anemia) AND (SARS-CoV-2 OR COVID-19 OR novel coronavirus).

Reference lists of the eligible studies will be screened manually for the identification of additional relevant reports.

### Inclusion and exclusion criteria

One author (Edouard Lansiaux) independently screened the articles by endnote software version 9 (https://endnote.com). The human observational studies, which reported the incidence of confirmed COVID-19 cases OR mortality rate OR severity assessment OR ICU admission among patients with beta-thalassemia minor, were included in this analysis. The language was limited to English. Studies such as case reports, review studies, and studies that did not have complete data for calculating incidences were excluded.

### Data extraction

The same author would individually extract the following data for each study (each author willcheck a separate database): title of the study, DOI, study design, timeline (retrospective or prospective), country, study type, age, gender, number of patients, number of patients with beta-thalassemic traits, number of COVID-19 patients, number of COVID-19 patients with beta-thalassemic trait, severity of COVID-19 (asymptomatic or mild, moderate, severe), mortality number, ICU admission, comorbidities [Diabete mellitus, Hypertension, Cardiovascular disease, Pulmonary hypertension, Cancer, Chronic Pulmonary Disease (emphysema or/and bronchitis), Autoimmune disease (Hashimoto thyroiditis, rheumatoid arthritis, systemic lupus and autoimmune hepatitis), Hypercholesterolemia, Stroke). Reference lists of the included articles will be screened for additional relevant studies.

Datasets will be extracted and managed on Microsoft Excel. For MS Excel extraction sheet, we would use the Cochrane Effective Practice and Organisation of Care (EPOC) Template modified for the needs of our study.

### Study quality assessment

The quality of the included studies was assessed by two independent assessors (Supplementary File 1). The JBI (Joanna Briggs Institute) critical appraisal checklist for cross-sectional and observational studies reporting incidence data. This checklist consists of nine items that evaluate the sampling process, data analysis process and statistical methods, study settings, measurement tools and response rate. The quality score of more than 5 was considered as moderate-high quality and included in meta-analysis. If there was any disagreement between assessors, it was resolved by discussion with authors.

### Statistical analysis

Data will be aggregated based on study design, location of reporting (country), participants demographic characteristics, etc.

Firstly, we will make a qualitative synthesis, describing the main characteristics of the studies. If possible based on data reported in extracted studies, meta-analyses will be performed on each of the included parameters using R and its package “meta”. Statistical significance will be set at P 0.05 unless stated otherwise in the study.

Concerning the meta-analysis first outcomes (such as the COVID-19 incidence, ICU admission incidence and mortality incidence estimations), they will be assessed with Freeman-Tukey, the variance estimation with a DerSimonian and Laird method, and the Cochran Q and I^2^ method to explore statistical heterogeneity at a significance value of 0.10. For studies with heterogeneity, we used a random effect model, while for those without significant heterogeneity, the fixed-effect model was applied [46]

Concerning the other main outcomes (severity odd-ratios), relative risks/risk ratios/hazard ratios for individual studies will be combined using a random-effects meta-analysis (based on type of variables - KM orCox regressional model). Each one will be assessed individually.

Moreover, pooled and individual data of the included surveys were depicted by Forest Plots. Publication bias was checked by the Egger test [47].

## Results

### Literature search and study characteristics

Figure 1 shows the PRISMA flowchart of the data selection process. The systematic search resulted in 2265 initial records, of which 871 were excluded as duplicates, 497 as irrelevant records due to the study date. 897 full-text articles were assessed for eligibility according to our inclusion criteria. Finally, 3 articles (3 studies) were found to be appropriate for quantitative synthesis [6,42,43] (Figure 2). The main characteristics of the studies are summarized in Table 1. The study designs mainly included cross-sectional methods. Publication bias was not significant according to the Egger’s test [15] (Table 2).

**Table 1.** Main characteristics of the included studies

|  |  |  |  |
| --- | --- | --- | --- |
| References | 10 | 11 | 6 |
| Timeline | Prospective | Retrospective | Prospective |
| Duration (days) | 61 days | 181 days | 30 days |
| Country | Greece | Greece | Italy |
| Study type | Cohort study | Cohort study | Case-control |
| Total patients | 255 | 760 | 801 |
| Total patients affected by COVID-19 | 255 | 760 | 182 |
| B-thal heterozygote patients | 45 | 189 | NR |
| B-thal heterozygote patients affected by COVID-19 | 45 | 189 | 19 |
| Severity |  |  |  |
| Total patients |  |  |  |
| Mild affected by COVID-19 | 68 | 190 | 143 |
| Moderate affected by COVID | 113 | 373 | 39 |
| Severe and Critical affected by COVID-19 | 74 | 197 |  |
| B-thal heterozygotes |  |  |  |
| Mild affected by COVID-19 | 5 | 15 | 19 |
| Moderate affected by COVID | 19 | 66 | 0 |
| Severe and Critical affected by COVID-19 | 21 | 56 |  |
| Mortality |  |  |  |
| Total deceased | 70 | 189 | NR |
| B-thal heterozygote deceased | 20 | 53 | NR |
| Total deceased by COVID-19 | 70 | 189 | NR |
| B-thal heterozygote deceased by COVID-19 | 20 | 53 | NR |
| ICU admission |  |  |  |
| Total ICU | 53 | NR | 39 |
| B-thal heterozygote ICU | 11 | NR | 0 |
| Comorbidities in general population |  |  |  |
| Male | 153 | 448 | 70 |
| Age: mean +/- SD | 61.56 (±16.597) | 62.21 (±16.42) | 53.2 +/- 18.1 |
| Diabete mellitus | 53 | 156 | 6 |
| Hypertension | 142 | 420 | 27 |
| Cardiovascular disease | 82 | 227 | NR |
| Cancer | 29 | 85 | 4 |
| Chronic Pulmonary Disease (emphysema or/and bronchitis) | 32 | 94 | 2 |
| Autoimmune disease (Hashimoto thyroiditis, rheumatoid arthritis, systemic lupus and autoimmune hepatitis) | NR | NR | 22 |
| Hypercholesterolemia | 113 | NR | 18 |
| Stroke | 50 | 142 | 14 |
| Comorbidities in B-thal minor population |  |  |  |
| Male | NR | NR | NR |
| Age: mean +/- SD | NR | NR | NR |
| Diabete mellitus | NR | NR | NR |
| Hypertension | NR | NR | NR |
| Cardiovascular disease | NR | NR | NR |
| Cancer | NR | NR | NR |
| Chronic Pulmonary Disease (emphysema or/and bronchitis) | NR | NR | NR |
| Autoimmune disease (Hashimoto thyroiditis, rheumatoid arthritis, systemic lupus and autoimmune hepatitis) | NR | NR | NR |
| Hypercholesterolemia | NR | NR | NR |
| Stroke | NR | NR | NR |

**Table 2.** Computed results concerning the different studied outcomes

|  |  | Incidence Rate<br>(95% CI) | Severity rate<br>(95% CI) | ICU admission rate<br>(95% CI) | Mortality rate<br>(95% CI) |
| --- | --- | --- | --- | --- | --- |
| Overall | index | 0.9250<br>(0.5752;1.4877) | 1.5933<br>(0.4884;5.1981) | 0.3620<br>(0.0025;51.6821) | 1.8542<br>(0.7819;4.3970) |
|  | z | -0.32 | 0.77 | -0.40 | 1.40 |
|  | p value | 0.7479 | 0.4400 | 0.6881 | 0.1610 |
| I<sup>2</sup>(%) |  | - | 90.0%<br>(73.1%-96.2%) | 91.7%<br>(71.0%-97.6%) | 85.1%<br>(39.7%;96.3%) |
| p value |  | - | 0.0001 | 0.0005 | 0.0095 |
| Egger's<br>test | 95 % CI<br>for bias | - | (-4.65;-0.12) | - | - |
|  | p-value | - | 0.3138047 | - | - |

**Figure 2.**
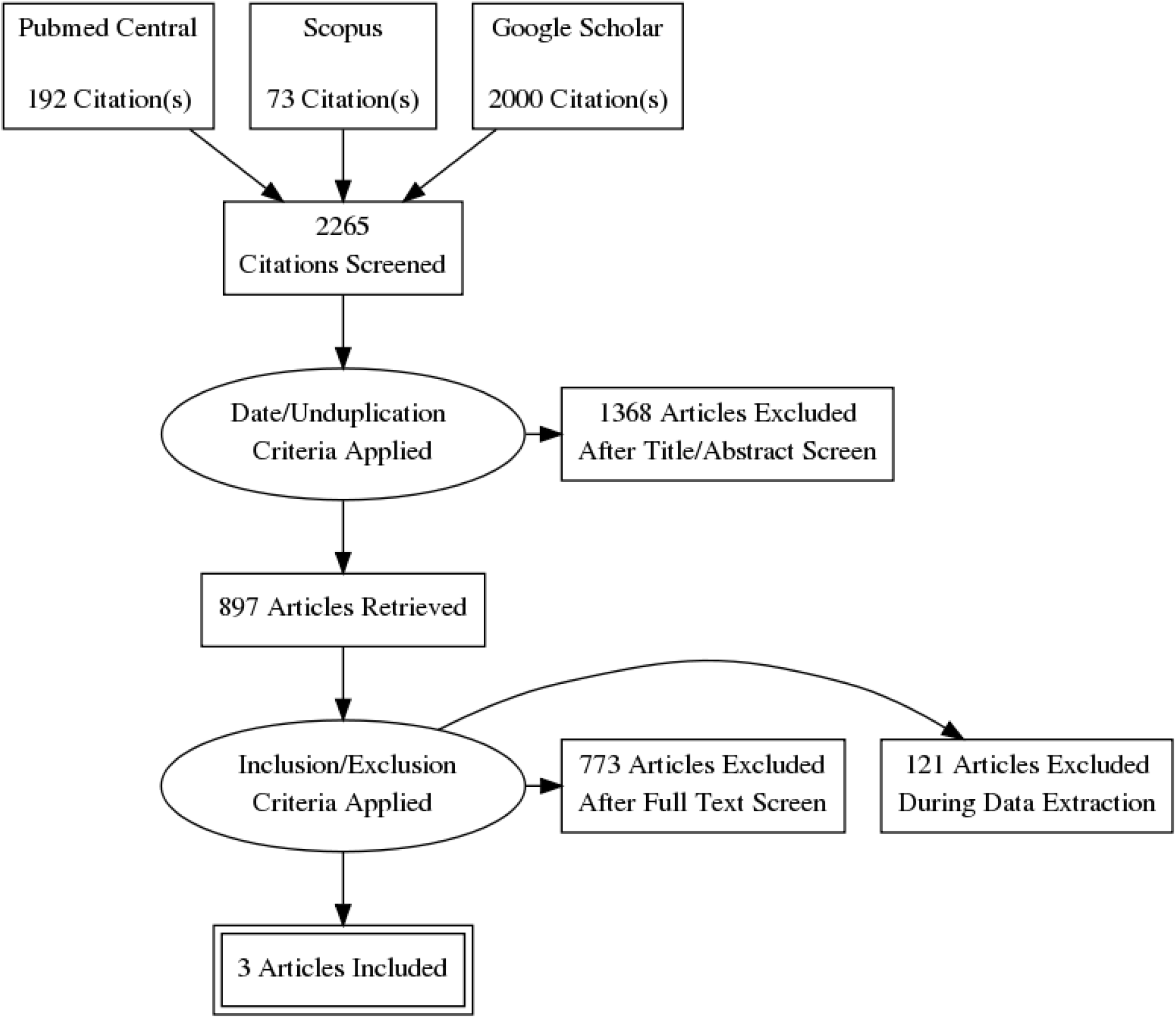
Flowchart of study identification and selection process

### Main outcomes

#### COVID-19 incidence

Only one study [6] was included into the analysis of the COVID-19 incidence into beta-thalassemic carriers. Therefore, we obtained a reduction of the incidence with an Incidence Rate Ratio of 0.9250 but it was not significant with its p-value of 0.7479 and the trust interval [0.5752;1.4877] (Figure 3).

**Figure 3.**
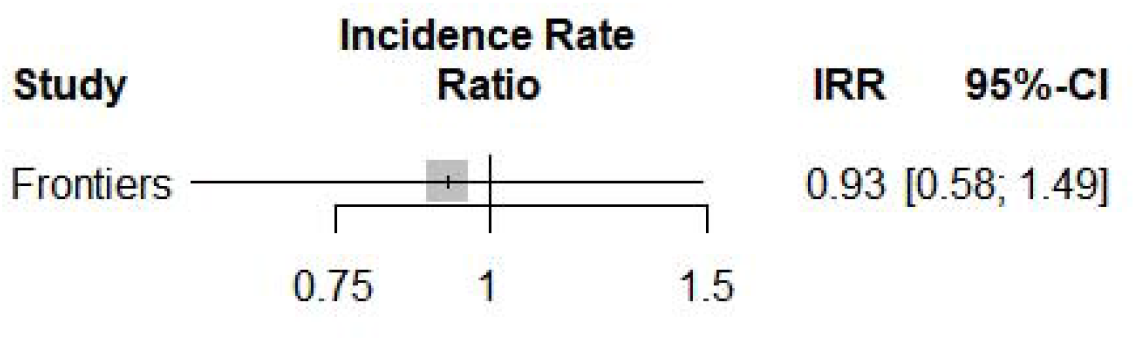
Incidence Rate Ratio of COVID-19 into beta-thalassemia carriers

#### COVID-19 severity Odds-Ratio

All of the three studies [6,42,43] were included here to compute an Odds-Ratio of 1.5933 which suggested an increase of severity into the beta-thalassemic carriers compared to the normal population. However, it stays insignificant with a 0.4400 p-value and a [0.4884;5.11981] trust interval. The random-model effect was used due to the heterogeneity (I^2^=90.0%) (Figure 4). Furthermore, selected studies have a large heterogeneity I^2^=91.7%(71.0%-97.6%) but didn’t indicate the presence of funnel plot asymmetry according to the Egger’s test (Table 2).

**Figure 4.**
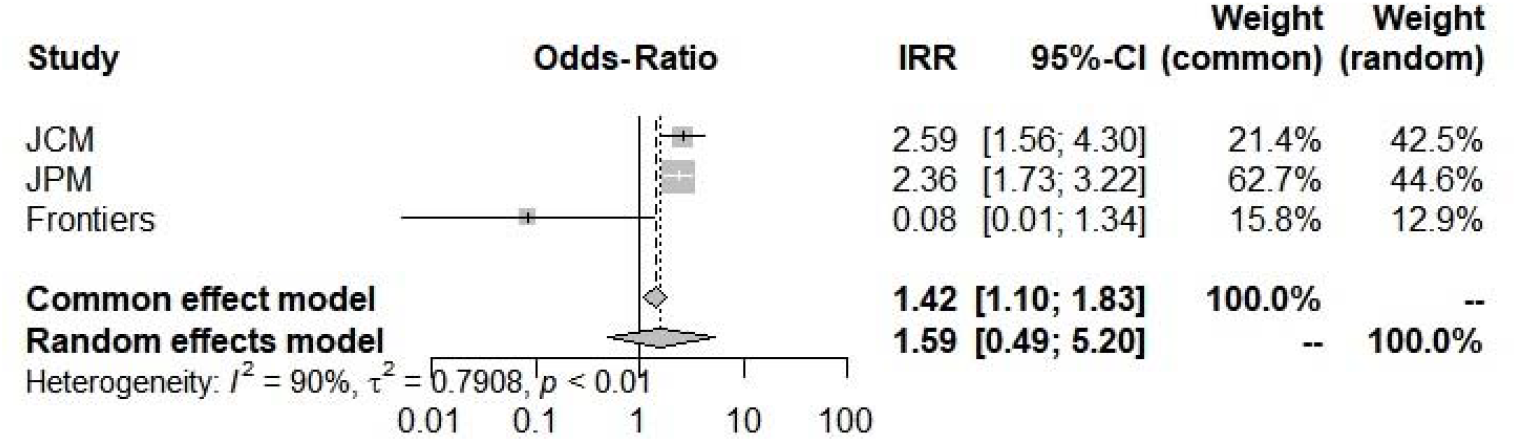
Odd-Ratios of COVID-19 severity into beta-thalassemia carriers

#### ICU admission incidence

Concerning ICU admission incidence, according to the random-effects model, we have computed an Incidence Rate Ratio of 0.3620(0.0025;51.6821) which suggests a large and insignificant decrease into the COVID-19 ICU admission concerning beta-thalassemia carriers compared to general population (Figure 5 and Table 2). Moreover, selected studies [6,42] have a broad heterogeneity I^2^=91.7%(71.0%-97.6%).

**Figure 5.**
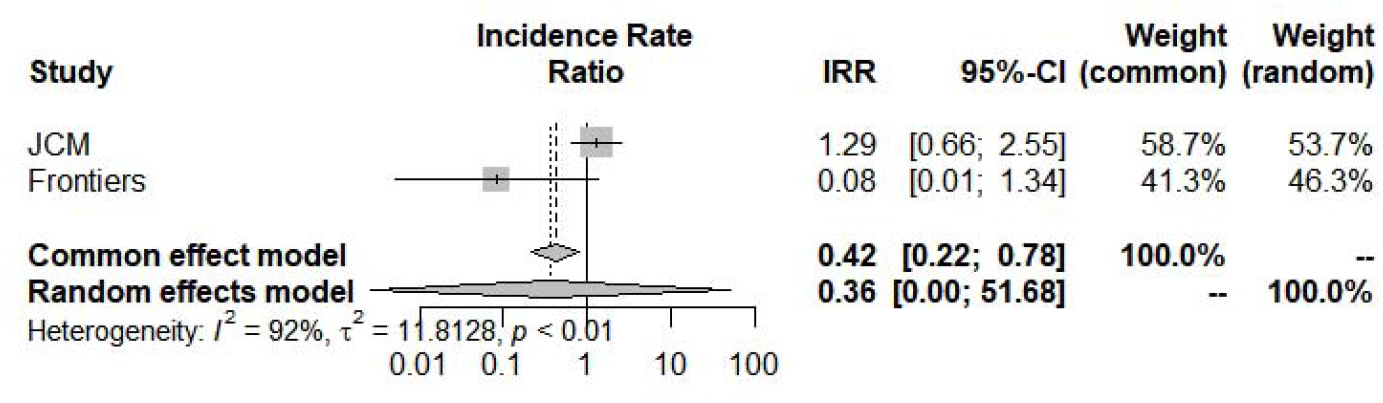
Incidence Rate Ratio of ICU admission for COVID-19 into beta-thalassemia carriers

#### Mortality incidence rate

The last outcome was the mortality incidence rate : beta-thalassemia carriers have shown a larger but insignificant propensity 1.8542(0.7819;4.3970) to die with a COVID-19 infection (Figure 6 and Table 2). Selected studies [42,43] have a broad heterogeneity I^2^=85.1%(39.7%;96.3%).

**Figure 6.**
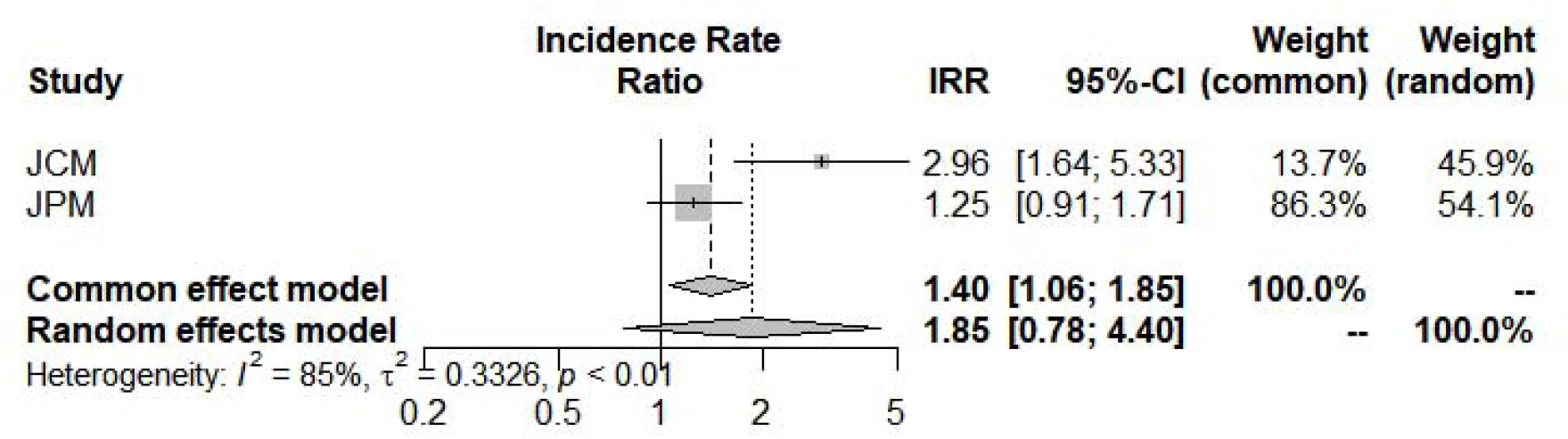
Mortality Incidence Rate Ratio for COVID-19 into beta-thalassemia carriers

## Discussion

According to our systematic-review meta-analysis, beta-thalassemia carriers could be lessCOVID-19 affected than general population [IRR= 0.9250(0.5752;1.4877)], affected by COVID-19 with a worst severity [OR=1.5933(0.4884;5.1981)], less admissible into ICU [0.3620(0.0025;51.6821)] and more susceptible to die from COVID-19 or one of its consequences [1.8542(0.7819;4.3970)]. However, all of those results stay insignificant with a bad p-value (respectively 0.7479, 0.4400, 0.6881, 0.1610).

This non-significance could be mainly explained by the small number of included studies (n=3). That’s our first limitation. The second consists into studies type; indeed, we couldn’t assess a real incidence based on only one case-control study and two others selected studies were COVID-19 positive cohorts.Therefore, more datasets are needed to be published in order to obtain significant incidence rate ratios and odds-ratios.

In spite of those terrible limits, this systematic-review meta-analysis opens the way to the confirmation of a possible protection/immunity of beta-thalassemia carriers against COVID-19 concerning incidence and ICU admission markers [7,8]. However, more epidemiological studies are required in the aim to obtain a strong signal.

## Data Availability

All data produced in the present work are contained in the manuscript

## Notes

<u>Transparency declaration</u>: All authors declare no support from any organization for the submitted work other than that described above; no financial relationships with any organizations that might have an interest in the submitted work in the previous 3 years; and no other relationships or activities that could appear to have influenced the submitted work.

### Competing Interest Statement

The authors have declared no competing interest.

### Funding Statement

This study did not receive any funding

### Author Declarations

The study used (or will use) ONLY openly available human data that were originally located at Pubmed central, Google Scholar and Scopus

### Summary of Updates

Adding more perspectives upon this physiopathological hypothesis

