## Supplementary Table 1 for "Beta-Thalassemia minor and SARS-CoV-2, prevalence, severity, morbidity and mortality: a systematic review study" (from another version of this preprint: version 1, 2022-07-25)

| References | 6 | 10 | 11 |
|---|---|---|---|
| Was the sample appropriate to address the target population? | yes | yes | yes |
| Were study participants sampled in an appropriate way? | yes | no | no |
| Was the sample size adequate? | yes | yes | yes |
| Were the study subjects and the setting described in detail? | yes | yes | yes |
| Was the data analysis done with sufficient coverage of the identified sample? | yes | yes | yes |
| Were valid methods used for the identification of the condition? | yes | yes | yes |
| Was the condition measured in a standard, reliable way for all participants? | yes | yes | yes |
| Was there an appropriate statistical analysis? | yes | yes | yes |
| Was the response rate adequate, and if not, was the low response rate managed appropriately? | yes | yes | yes |
| Total score | 9 | 7 | 7 |

Supplementary Table 1. Included studies quality assessment according to Joanna Briggs Institute critical appraisal checklist
